# Proximity and metabolic activity of the Tumour Microenvironment as predictors of survival in High Grade Serous Ovarian Cancer (HGSOC)

**DOI:** 10.1101/2025.03.14.25324006

**Authors:** Naomi Berrell, Aaron Kilgallon, Meg L Donovan, Clara Lawler, Chin Wee Tan, Kidane S Embaye, Rafael Tubelleza, James Monkman, John F Fraser, Ken O’Byrne, Ruby Yun-Ju Huang, Arutha Kulasinghe

## Abstract

High grade serous ovarian cancer (HGSOC) is a lethal gynaecological malignancy, most often detected at the late stages of disease, and at which point, response to therapy is only seen in a minority of patients. As new treatments are introduced, there is a growing need to better understand how the tumour microenvironment (TME) is predictive of therapeutic benefit. Here, we spatially analysed tumour samples from 55 HGSOC patients using high-plex spatial proteomics to characterise the TME and define the contributions to patient survival in cisplatin resistance. Using a custom-developed 48-plex cyclic immunofluorescent protein panel, we analysed tumour and immune cell interactions and their functional and metabolic profiles. We found that a higher number of CD66+ cells in a 50µm radius of tumour cells (p = 0.025) and a higher number of cytotoxic CD8 T -Cells within a 10 µm radius of the tumour boundary were associated with improved overall survival (p = 0.03). We found that in areas of metabolically active tumour cells, the proximity of T-Reg Cells was associated with better overall survival when the activity of the tumour was increased in either citrate synthase activity (p = 0.0064) or high nitric oxide (p = 0.017). However, with metabolically less active tumours, there was an association with worse overall survival (p = 0.033). Taken together, our data suggest that a comprehensive functional understanding of the TME is needed to quantify the therapeutic benefits of immunotherapy in HGSOC.

## BACKGROUND

Globally, ovarian cancer (OC) has an incidence of over 313,000 cases and results in over 207,000 deaths per year (GLOBOCAN 2020)^1^. Symptoms, such as menstrual cramps, are generally mild, and coupled with unreliable screening tools patients often receive a late-stage diagnosis with poor treatment outcomes^2^. Prognoses for patients with advanced disease stages are usually poor, with a five-year survival of less than 17%^3^. When detected early, most patients will initially recover by radical surgery and/or adjuvant chemotherapy but can experience reoccurrence^2,4,5^. At later stages of disease, however, surgery is insufficient to clear microscopic lesions and treatment success relies on the efficacy of chemotherapy agents^2^. With limited systemic and/or targeted therapies, it is becoming increasingly important to understand the properties of the underlying TME which may provide insights into more effective therapeutic strategies.

As therapy resistance in this tumour type is common, there is an unmet clinical need for improved treatments targeting OC during late-stage disease, as well as biomarkers to accurately predict a patient’s response to therapies^6^. Altered cellular energetics stands as one of the hallmarks of cancer and contributes to therapy resistance and immune evasion. Over several decades, many studies have revealed that the alteration to various metabolic pathways, such as amino acid metabolism, increased glycolysis, or fatty acid metabolism can contribute to tumour proliferation, therapy resistance and immunosuppressive tumour microenvironments^7–9^ ^10,11^.

Results from a number of pre-clinical and clinical studies suggest that the exploitation of these dysregulated pathways and metabolic addictions may improve the effectiveness of current and future therapeutics ^12^ ^13,14^.

Spatial proteomics allows for insights into the TME by a comprehensive profiling of the cellular, tissue, and organ immune contexture^6^. Spatial proteomics facilitates the analysis of many biomarkers across the TME and in the tumour and stroma simultaneously, enabling us to profile the tumour-immune composition and cell-cell interactions in tissues at multiple levels of complexity; including capturing cell types and their functional and metabolic states^6,15^. Targeting antibodies for enzymes associated with metabolic activity allows for enhanced profiling of dysregulated metabolic pathways within the TME. Moreover, these approaches allow for sub-phenotyping of the individual cell types using functional characterisation. This approach enables the development of large, targeted panel proteomic profiles in a non-tissue-destructive and high-throughput manner for translational cancer research.

In this study, we present a spatial proteomic study of the TME of 55 OC patients using a custom-developed 48-plex mIF protein panel. Our panel encapsulates the major hallmarks of cancer and spatially characterises HGSOC tumour and immune cells and their functional and metabolic states. The spatially resolved HGSOC TME profiles were measured against clinical endpoint of overall survival (OS). These analyses include identifying cellular neighbourhoods common across the cohort, highlighting proximity density trends between two distinct cell types, profiling cells located at the tumour boundary, and characterising spatial patterns occurring within the tumour that associate with OS.

## METHODS

### Study Design

This study profiled formalin-fixed paraffin-embedded (FFPE) tissue microarray (TMA) consisting of single 1mm diameter tissue cores from 55 women with HGSOC (TA4359, SKU: 69574359, Tristar Technologies, USA). The tissues consisted of samples collected between 2009 and 2017 with informed written consent provided by the collaborating hospital sites. The tissue samples were collected post-therapy from various regions (peritoneum, ovaries, omentum, pelvic mass, and inguinal lymph node) and were predominantly composed of the high grade serous histological subtype (n = 51). However, samples also included two low grade serous carcinomas, one clear cell carcinoma and one mixed cell carcinoma. The downstream analysis was only conducted on the high grade serious histological diagnosis samples. After quality control, three additional samples were excluded due to poor tissue quality or staining artifacts. Apart from two stage-2 patients, all patients had advanced disease at diagnosis (stage 3 or higher). Patients received chemotherapy (1-4 rounds of Carboplatin/Paclitaxel (n=42), 3-6 rounds of single agent carboplatin (n=10), or unspecified treatment (n=3) before undergoing interval debulking surgery. However, information regarding treatment received in the post-surgical setting was not available. This study has University of Queensland Human Research Ethics Approval (2021/HE001936).

### Tissue Profiling

The TMA was subject to high-plex cyclic-IF using the PhenoCycler Fusion (Akoya Biosciences, USA), following established protocols as previously described^16^. A panel of 48 antibodies was custom-designed using the Hallmark Features of Cancer^17^, including markers for cell typing, cell functionality, and markers of cellular metabolism^15^ (Table 2). Briefly, FFPE tissue sections were baked at 60°C for 1 hour before being submerged in Histochoice Clearing Agent (VWR) to remove wax. Sections were then hydrated using decreasing concentrations of ethanol, followed by heat antigen retrieval in AR9 buffer (AR9001KT) using the standard settings of a conventional pressure cooker for 20 min. Following this, sections were washed in ddH_2_O hydration buffer and staining buffer solutions before staining with the customised antibody cocktail and allowed to incubate overnight at 4°C in a humidity chamber. H&Es images were reviewed by a histopathologist independent of the mIF cell typing. Cell typing annotations were checked against this ground truth annotations for tumour, stroma, and immune cells.

**Table 1:** Patient characteristics.

|  |  |
| --- | --- |
| **Histological diagnosis** |  |
| High grade serous | 51 |
| Low grade serous* | 2 |
| Mixed cell* | 1 |
| Clear cell* | 1 |
| **Adjuvant chemotherapy regime 1** |  |
| Carboplatin/Paclitaxel | 41 |
| Carboplatin | 9 |
| Unspecified | 5 |
| **Overall Survival** |  |
| Alive | 11 |
| Deceased | 44 |
| Overall survival (median) | 862 days (Range 172 - 2691 days) |
| **Stage at diagnosis** |  |
| 2a | 2 |
| 3 | 1 |
| 3c | 35 |
| 4 | 16 |
| 4b | 1 |
| **Site of Diagnosis** |  |
| Ovaries | 26 |
| Omentum | 13 |
| Ovary/peritoneal | 9 |
| Peritoneum | 4 |
| Inguinal lymph node | 1 |
| Pelvic mass | 1 |
| Unknown | 1 |
| **Age** |  |
| Median | 65 (Range 47 - 86) |

\* (was not used in analysis)

**Table 2:** Panel of markers.

| **Immune** | **Functional** | **Tumour** | **Metabolic** |
| --- | --- | --- | --- |
| CD107a | Cleaved Parp | Pan-Cytokeratin | CPT1A |
| CD11c | BCLXL | E-Cadherin (+ Structural) | GLUT1 |
| CD14 | CD38 |  | iNOS |
| CD20 | CD39 |  | ASCT2 |
| CD21 | FOXP3 |  | TCA (Citrate synthase) |
| CD4 | Granzyme-B |  | G6PD |
| CD66 | HLA-DR |  | pNRFR2 |
| CD68 | ICOS |  |  |
| CD141 | IDO1 |  |  |
| CD3e | IFN-g |  |  |
| CD45 | Ki67 |  |  |
| CD45RO | LAG3 |  |  |
| CD79 | MMP9 |  |  |
| CD8 | PD1 |  |  |
| CD56 | PDL1 |  |  |
|  | s100a |  |  |
|  | Tbet |  |  |
|  | TCF1 |  |  |
|  | VISTA |  |  |

### Cell segmentation and data QC

Qptiff files were imported into QuPath for visualisation, initial analysis, and quality control^18^. Stardist was used to perform cell segmentation based on nuclei staining^19^. A pixel classifier was user-specified using PanCk^+^ and PanCk^−^ regions to define tumour and stromal regions, respectively. This approach allows for the interrogation of tumoural and stromal markers within the TME. Quality control of antibody staining was performed by manual inspection of marker specificity, with markers displaying ubiquitous and non-specific staining removed from the analysis, as previously described^20^. The median expression of each marker for each cell was exported as a csv file, containing each cell’s unique object identifier, x-y coordinates, cell morphology features and core ID.

### Cell typing analysis

Using a subset of cell markers (CD44, CD68, CD3e, CD45, CD56, E-cadherin, Ki67, PanCK, CD20, CD14, CD8, SMA, nuclear size and nuclear diameter), unsupervised clustering was performed on Phenograph^21^cluster cell groups to assign tumour and non-tumour phenotypes. This marker subset were normalized using a percentile transformation prior to cell typing. Generated cell type annotations were then joined to the original AnnData structure that contained the raw data. For further phenotype classifications, when data were subset from the raw file, the same normalisation method was applied. Unsupervised clustering was initially performed over a range of resolutions and nearest neighbours but generally failed to generate meaningful clusters, necessitating supervised cell typing. To characterise cells further, the non-tumour cell phenotypes were classified using the gating method, mmochi^22^, that is based on a gaussian mixed model (GMM) combined with a user-defined hierarchy (Figure 2). Iteratively, cell types were imported back to QuPath for visualisation and threshold cutoffs manually adjusted to reflect true cell types. Broad cell types were assigned by interplay between cell-type heat maps and visualization of cell types on the raw image. Relevant functional markers were chosen to better define subsets of T cells, B cells, macrophages and tumour cells (Figure 2). Using a GMM, positivity for relevant markers was assigned to identify functional cell types. Tumour cells were phenotyped for functional markers and metabolic markers by the GMM and unsupervised clustering methods, respectively. For functional markers, tumours were labelled to be positive or negative for iNOS, IDO1, CD44, and IFN-g based on the GMM positivity classification. This approach allowed us to focus on specific functional interactions between immune and tumour cells. For the metabolic phenotyping, we opted to use an unsupervised clustering method to annotate these cells as we wanted to assess tumours in their whole metabolic state, which could include having multiple dysregulated metabolic pathways^21^. Data were subset to include the CPT1A, pNRFR2, ASCT2, Citrate-Synthase, G6PD, GLUT1 and iNOS markers, and unsupervised clustering was performed using the Leiden algorithm. A search over a range of *k* and *r* parameters was performed using a GPU-accelerated adaption of the Phenograph method from Scanpy to identify optimal parameters^21^. At a k-nearest neighbours value of 20 and Leiden resolution of 0.25, cells separated into distinct tumour phenotypes. Groups were then annotated based on the expression profiles displayed on a heatmap.

### Neighbourhood analysis

Cellular Neighbourhoods were created using a K Nearest-Neighbours (KNN) approach by specifying the number of neighbours reported per cell and the number of neighbourhoods to define ecosystems that have similar cells present near each other ^23^. Three neighbourhoods were defined using the tumour/non-tumour annotations to create tumour-dense, stromal-dense and mixed neighbourhoods (immune infiltrated tumour regions or tumour interface zones). The SPACEc bioinformatics package was used to generate more complex neighbourhoods across the cohort^24^. To determine an optimal number of neighbourhoods, a Distortion Score Elbow for miniBatchK-means was calculated, where the distortion score over a range of 20 neighbourhoods was determined to identify the elbow point.

### Proximity analysis

Using cell-type and spatial information, we computed various spatial metrics to compute relationships between sets of cells that could be associated with disease outcome. Using the Scimap package, we performed spatial density and volume analyses^25^. Proximity density and volume metrics from the Scimap package were calculated between cell type combinations for broad and functionally annotated cell types^25^. The proximity density measure estimates how often two cell types co-occurred within a pre-determined radius, divided by the total number of those cell types. Higher scores indicate that these cell types were more frequently associated with one another. The proximity volume metric evaluates how often two cell types co-occur within a radius relative to the total number of cells within the KNN subgraph. Proximity density and volume scores were calculated per tumour core over a range of distances between 5 and 100 µm by increments of 5 µm^24^. Scores were calculated using the area under the curve and were then compared by survival time. Using SPACEc, patch analyses were performed around specified neighbourhoods for each tissue core^24^. Edges of cell clusters were determined, followed by identification of cells within radii of 5 - 25 µm^24^. The proportions of cells in each radius were compared across survival groups and survival times.

### Validation using publicly available datasets

The Xena Browser, provided by the University of California Santa Cruz, was used to validate statistically significant findings from cell count and proportion analyses^26^. This interactive web-based tool allows users to assess publicly available datasets (including RNA, protein and spatial datasets) and perform statistical analysis based on overall survival or other available clinical information. Using data from the GDC TCGA Ovarian Cancer RNA-seq dataset, the gene expression from 425 patients was assessed by applying insights gained from evaluating hazard ratios on overall survival estimated by a Kaplan Meier analysis.

### Statistical analysis

This study utilised several statistical methods to identify statistically significant features that correlated with survival. To identify whether features are associated with overall survival, a univariate survival analysis was performed using a Kaplan Meier estimate between upper and lower quartiles and median cut-offs^27^. Statistical significance was estimated from a log-rank test using the Lifelines package^27^. Additionally, using the Lifelines package^27^, a multi-variate Cox-Proportional Hazard model was evaluated to compare the effect of multiple variables on patient outcome. To identify features that may be associated with survival over three years, the average findings for each patient were compared using the non-parametric Mann-Whitney U-test. Statistical significance was considered using unadjusted p-values where p < 0.05.

The stability selection method was implemented in the sharp R package with CoxPH models pulled from the glmnet library. The LASSO l1-regularized fits of CoxPH models to the cohort survival data selected 86 features at the optimal stability point of the model, and a final l1-weighted CoxPH model reduced this feature set to 52 features of interest with non-zero coefficients. Model parameters were sampled on a grid of threshold selection frequencies from 0 to 0.5. The final regularized CoxPH model to reduce the feature space was fit with the minimal regularization parameter (A = 0.03) suitable for a stable fit to the OS clinical data.

## Results

### The OC TME is characterised by a complex immunological profile

After quality control of tissues and staining quality, 5 cores were excluded from the analysis. Overall survival and status were varied across the cohort, with no obvious trends between tumour location (Figure 1A). The tumour cores were collected from the ovaries, peritoneum, inguinal lymph node, omentum and pelvic mass (Figure 1A). The characterisation of the OC TME was performed by a combination of unsupervised Phenograph clustering and supervised clustering to provide an accurate and biologically driven profile of immune-functional and metabolic markers based on cell types^21^. Tumour and non-tumour cells were separated using Phenograph to perform unsupervised clustering based on a subset of broad tumour and non-tumour phenotype markers^28^. Primarily, PanCK was used as a tumour marker to differentiate malignant cells from immune and other non-tumour cells. Using a subset of immune cell markers, we aimed to classify non-tumour types into broad immune phenotypes. The immune phenotypes included B cells, CD4^+^ T cells, CD8^+^ T cells, Double Negative (DN) (CD4^-^CD8^-^) T cells, fibroblasts, CD66^+^ cells, monocytic, structural cells, and blood vessels (Figure 1 B & D). The number and type of cells present within each core were heterogeneous. Most cores largely consisted of tumour and structural cells. CD8^+^ T cells, CD4^+^ T cells, monocytic cells and tumour cells were all subject to functional classification. Based on the expression of certain combination of the above markers, these cells were functionally annotated (Figure 1G). Overall, we observed 18 immune phenotypes after functional annotation, with tumours also annotated to be positive or negative for iNOS, Ki67, CD44, IFN-g, and IDO1 (Figure 2F-H). For the metabolic tumour phenotyping, 33 groups were generated based on the expression of CPT1A, pNRFR2, ASCT2, Citrate-Synthase, G6PD, GLUT1, and iNOS, but was reduced to 9 main phenotypes (Figure 1C & E). iNOS was used across both the functional and metabolic phenotypes due to its contribution to both functional and metabolic states.

**Figure 1:**
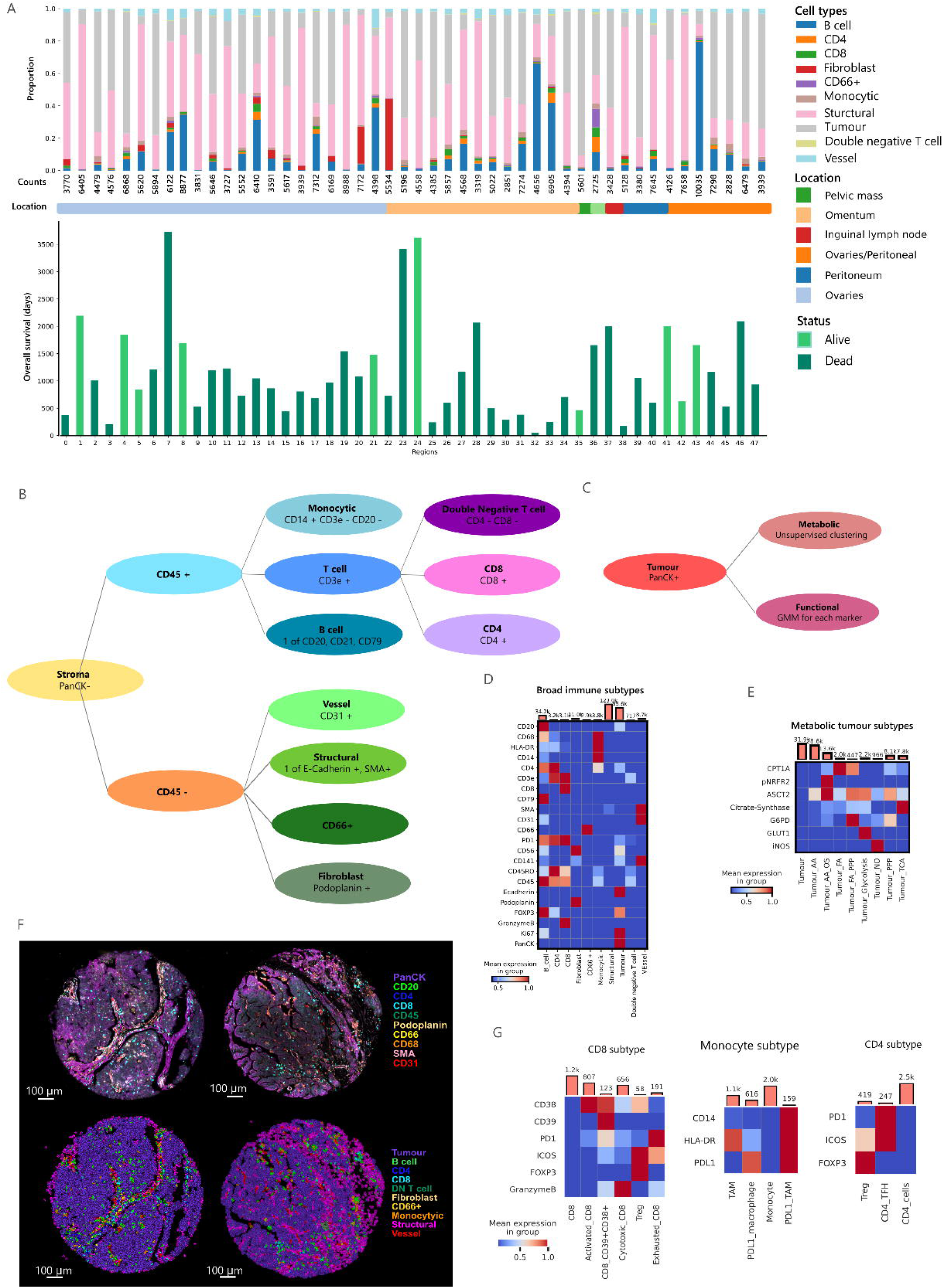
**A)** Patient cohort information. **Top:** Bar graph showing the total cell counts and the cellular proportions across the samples included in this analysis. **Middle:** Location of tissue biopsy. **Bottom:** Overall survival in days for each patient. Bars are coloured based on survival status, light green = alive, dark green = dead. **B)** For non-tumour cells, hierarchical cell typing was performed following the tree diagram shown. Non-tumour cells were classified based on negative PanCK expression. **C)** Tumour cells (PanCK positive cells) were further phenotyped using unsupervised clustering for metabolic markers, (**E)** and were also annotated based on positive expression of functional markers using a Gaussian Mixture Model (GMM). **D)** Heatmaps representing cellular phenotypes and their mean relative expression of markers. **F)** Representative images of tumour cores, with immunofluorescent images (above) shown next to the cell annotations mapped onto the tumour cores (below). **G)** Immune subtypes were further annotated using a GMM classifier. Cells were annotated based on the combinations of markers expressed. Heatmaps represent the cell phenotypes and relevant markers used for phenotypes.

### Spatial metrics

Neighbourhoods were generated based on functionally annotated immune cells, where 5 neighbourhoods were selected from the elbow point found using a distortion plot (Supplementary Figure 1). Neighbourhoods were annotated based on abundance of cell types using a heatmap and proportions bar chart (Figure 2 A-B). Mapping of neighbourhoods onto tumour samples was done to visualise the differences among neighbourhoods in-situ (Figure 2C). Based on the proportion of neighbourhoods in each sample, only the immune low tumour neighbourhood was significantly associated with overall survival, as per the Kaplan Meier (KM) analysis (p = 0.0371), whereas a higher proportion of these tumour neighbourhoods was associated with poor survival (Figure 2D).

**Figure 2:**
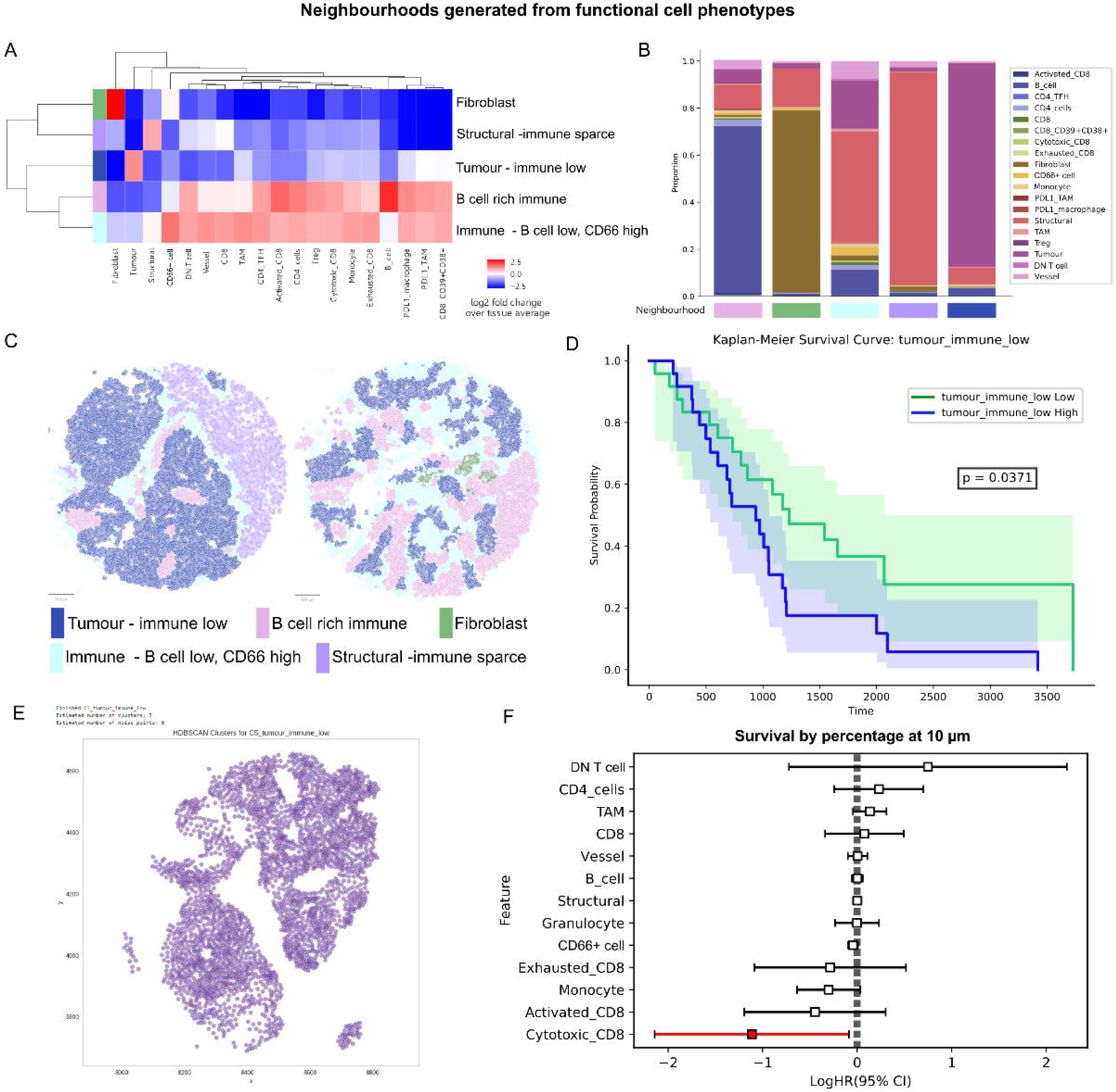
**A)** Heatmap showing relative abundance of functional immune phenotypes within cellular neighbourhoods. **B)** Bar graph showing proportions of cell types in each neighbourhood. **C)** Visualisation of cellular neighbourhoods mapped onto tissue cores. **D)** Kaplan Meier analysis based on proportion of tumour immune-low neighbourhoods are correlated with overall survival. **E)** Patch analysis identifies patches of tumour immune-low neighbourhoods, where the boundary of these patches is then used to measure cellular abundance in specified radius around these patches. **F)** A fitted Cox Proportional Hazard model of cell types proportions within 10 microns from the tumour immune low boundary. Features that are statistical significance **are** highlighted in red.

SPACEc patch analysis identifies clusters of one cell type or neighbourhood and determines the cellular proportions present within user defined radii surrounding the outside of these patches^24^. We performed patch analysis around clusters of the immune low tumour neighbourhoods to identify differences in the proportions of surrounding cells between patient groups (Figure 2E). Proportions of cell types were calculated at distances of 5 µm, 10 µm, 15 µm, 20 µm, and 25 µm. For each distance, we performed a CoxPH model fit to identify cells within each radius that might be contributing to patient outcomes. Cells were excluded from the model if the variance of an outcome group was less than 0.01. At 5 µm and 25 µm, there were no statistically significant findings. At 10µm radius, cytotoxic CD8 T cells were enriched in patients with hazard ratio (HR) <1, indicating association with improved patient outcomes (p = 0.03) (Figure 2F). At 15 µm, tumour associated macrophages (TAMs) were identified to have an HR >1 (p = 0.04) (Supplementary figure 2B) and at 20 µm, double negative T cells had an HR> 1 (p = 0.04), whereas B cells (p = 0.05) and monocytes (p = 0.02) had HR’s <1 (Supplementary figure 2 C-D).

Proximity density analysis was performed to identify trends that were occurring within the tissue that suggest common cellular distributions between tumour cells and other cells within the TME and whether these were associated with survival outcomes (Figure 4A). The same method was employed for cell type combinations at a distance of 50 µm, and over a range of distances 25 - 100µm in 5µm increments, where the area under the curve (AUC) was used to assess the distribution of cells. At 50 µm, the CoxPH model of cells locations in relation to tumour cells showed that exhausted CD8+ T cells (p = 0.05), fibroblasts (p = 0.035) and PDL1+ TAMs (p = 0.0077) were associated with worse survival (HR>1), whereas increased proximity density between CD66+ cells (p = 0.017) or activated CD8+ T cells (p = 0.021) were associated with improved outcomes (HR<1) (Figure 3B). KM analysis of these cell types revealed only the proximity density scores between tumour cells and PD-L1+ TAMs (p = 0.0123) (Supplementary figure 3B) and CD66+ cells (p = 0.0254) (Figure 3C) were statistically significant. Higher proximity density scores between PDL1+ TAMs and CD8+ T cells (p = 0.018), exhausted CD8+ T cells (p = 0.023), cytotoxic CD8 T cells (0.0495) and activated CD8 T cells (p = 0.034) were all associated with worse OS (Supplementary figure 4). Furthermore, the KM analysis modelled on the proportions of both PDL1+ TAMs (p = 0.0168) and CD66+ cells (p = 0.0151) at a whole core level were also statistically significant (Supplementary Figure 3A & C). Higher proximity density scores between CD66+ cells and structural cells (p = 0.019) were also associated with improved outcomes (Supplementary figure 3D). Over multiple radii, we see that patients alive past two years tend to consistently have higher proximity density score between tumour and CD66+ cells compared to patients who died prior to two years (Figure 3D). Although not statistically significant, there was a trend that a greater AUC was associated with better survival (p = 0.0867) (Supplementary figure 5).

**Figure 3:**
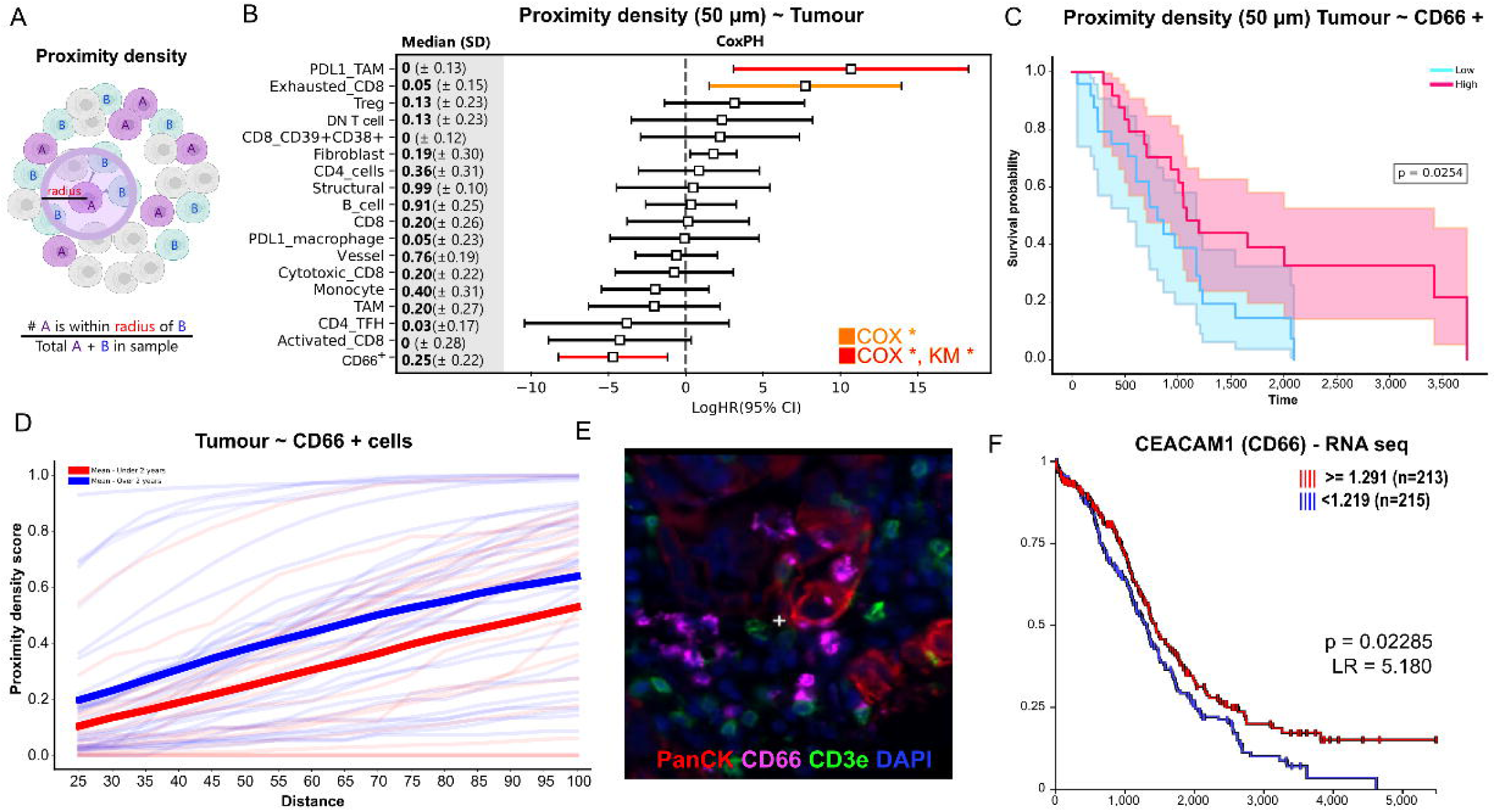
A profile of proximity density scores of cell types in our ovarian cancer dataset. **A)** The proximity density algorithm calculates the ratio of two cells within a pre-defined radius and the total number of those two cell types present in each sample. **B)** Proximity density scores for cell combinations featuring tumour cells (mean +/- SD) are shown along the left column. Cox proportional hazards (CoxPH) model of proximity density scores (based on 50 µm radius) between tumour and immune cells. Statistical significantly combinations for both CoxPH and Kaplan Meier (KM) models are highlighted in red, and statistical significance exhibited in only CoxPH and not KM models are highlighted in orange. **C)** KM analysis of proximity density scores between tumour and CD66+ cells show an association with improved survival (based on median proximity density, with higher scores shown in orange and lower scores shown in blue). **D)** Proximity density scores between tumour and CD66+ cells over a range of distances between 25 µm and 100µm. Blue lines represent patients who died prior to two years, red represents patients who lived past two years. The mean scores for each survival group is shown bolded in their respective colours. **E)** A representative immunofluorescent image of CD66+ cells in a tumour region. **F)** External validation via analysis of publicly available bulk RNA-seq data from the GDC TCGA Ovarian cancer dataset showed that high CEACAM1(CD66) expression is associated with improved survival (red), whereas lower expression is associated with poorer survival (blue line). Data was analysed using Xenabrowser ^29^.

**Figure 4:**
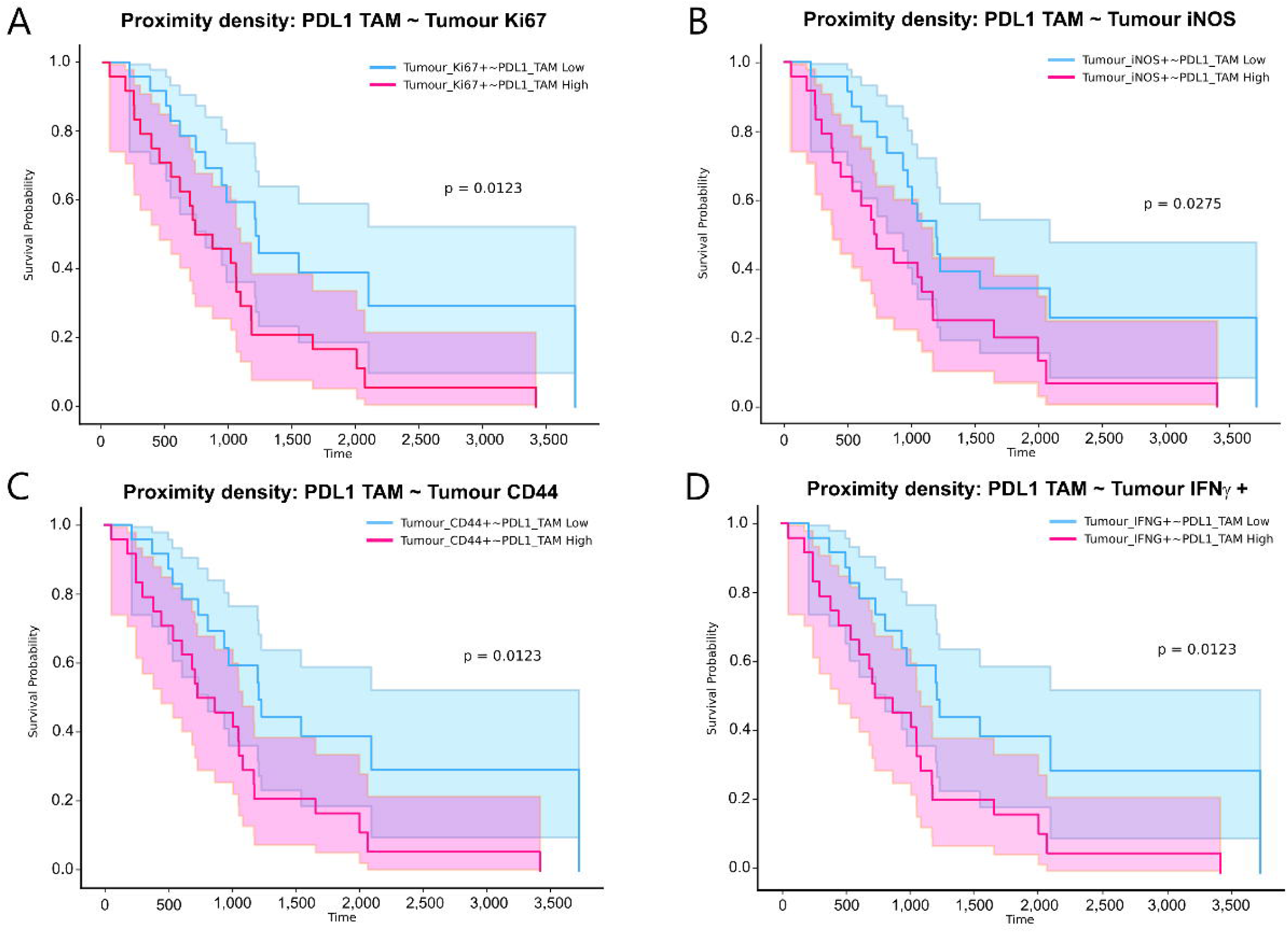
**A-C)** Proximity density scores between tumour cells with a metabolically low (**A**), tricarboxylic acid cycle (TCA) high (characterised by high citrate synthase expression) **(B)** or nitric oxide (NO) high phenotype (characterised by high inducible nitric oxide synthase expression) **(C).** Median values are shown as an orange line. Hazard ratios from a fitted Cox Proportional Hazards (CoxPH) model of proximity density score features, based on occurrences with a 50 µm radius between different metabolic phenotypes of tumour cells and immune cells. Red lines represent statistical significance in both CoxPH and Kaplan Meier models. Orange lines represent statistical significance in the CoxPH model. **D)** Hazard ratios from a CoxPH model based on tumour cells with low/no expression of metabolic markers. **E)** Hazard ratios from a CoxPH model in relation to TCA high tumour cells. **F)** Hazard ratios from a CoxPH model in relation to NO high tumour cells.

CD66 staining was localised to a subset of cells that did not show PanCK and CD3e staining (Figure 3E). To cross-validate the presence of CD66 positivity that was found in our non-spatially focused analysis, we analysed CD66 (CEACAM1) expression within the GDC TCGA Ovarian Cancer bulk RNA expression (n=429) dataset. Given that CD66 was the only marker expressed on this subset of cells, we then assessed the univariate expression of CEACAM1 (CD66a), and it was found that higher expression of CEACAM1 was associated with improved survival (p = 0.023) (Figure 3F).

### The tumour-metabolic profile

The metabolic profile of tumour cells is known to alter the TME and can lead to impaired immune cell function, immune evasion, and drug resistance. Proximity density analysis was performed at a 50 µm radius to identify interactions between immune and tumour cells based on metabolic dysregulation. Tumour cells that did not highly express any metabolic markers in our panel were classed as ‘metabolic low’. A proximity density analysis was performed between the metabolically distinct tumour phenotypes and immune cells (Figure 4A-C). Hazard ratios from fitted CoxPH models were calculated for each tumour subtype and immune cells (Supplementary figure 6). For the metabolically low tumours, we identified these immune cells that associated with worse outcome (HR>1): PDL1+ TAMs (p = 0.0062), Tregs (p = 0.033) and Fibroblasts (p = 0.0060) (Figure 4D).

Proximity density scores between tricarboxylic acid (TCA) high tumours and immune cells found that high proximity density scores with Tregs were associated with improved survival (p = 0.0064), however a high proximity density score with PDL1+ TAMs correlated with worse survival (p = 0.0036) (Figure 4E). For this tumour type, statistically significant associations existed between tumour cells and vessel cells (p = 0.027) and activated CD8 T cells (p = 0.020), predicting worse survival, and PDL1+ macrophages (p = 0.026) were associated with improved outcomes (Figure 4E). Nitric oxide (NO) high tumours also identified T-Reg Cells to be associated with improved outcomes (p = 0.017), however PDL1+ TAMs were not associated with survival outcomes (p = 0.73) for this tumour subtype. While in metabolically low and TCA high tumours, cytotoxic CD8+ T cells did not associate with survival outcomes. However, in the NO comparison, these cytotoxic CD8+ T cells had a HR < 1 (p = 0.021) (Figure 4F).

### Functional markers expressed on tumour cells are associated with impaired immune response

When assessing interactions between tumour and immune cells, tumour cells were functionally annotated to be either positive or negative for the IDO1, iNOS, Ki67, CD44 and IFN-g functional markers using a GMM classifier. Based on these annotations, we assessed patterns of immune cells in proximity to any of these functional markers and whether these correlated with survival outcomes. Using the proximity density score, we identified several patterns consistently occurring across multiple immune types, when in proximity to functional tumour phenotypes, that were identified to be significantly associated with overall survival. High proximity density scores between PDL1+ TAMs and CD44+ (p = 0.0123), Ki67+ (p = 0.0123), iNOS+ (p = 0.0275), and IFNG+ (p = 0.0123) tumour cells were associated with worse outcomes (Figure 5). The same relationship was seen for Ki67+ (p = 0.0010), iNOS+ (p = 0.0291), IFNG+ (p = 0.0283) and IDO1+ (p = 0.0150) tumour cells with Fibroblasts (Supplementary Figure 7A-D). High proximity density scores between PD1+ CD8 T cells and either Ki67+ tumour cells (p = 0.0418) or IDO1+ tumour cells (p = 0.0487) were found in patients who had shortened overall survival (Supplementary figure 7G-H). This was also seen between CD8+ T cells and IDO1+ (p = 0.021), Ki67+ (p = 0.0478), and iNOS+ tumour cells (p = 0.0034) (Supplementary Figure 7E-F, I).

**Figure 5:**
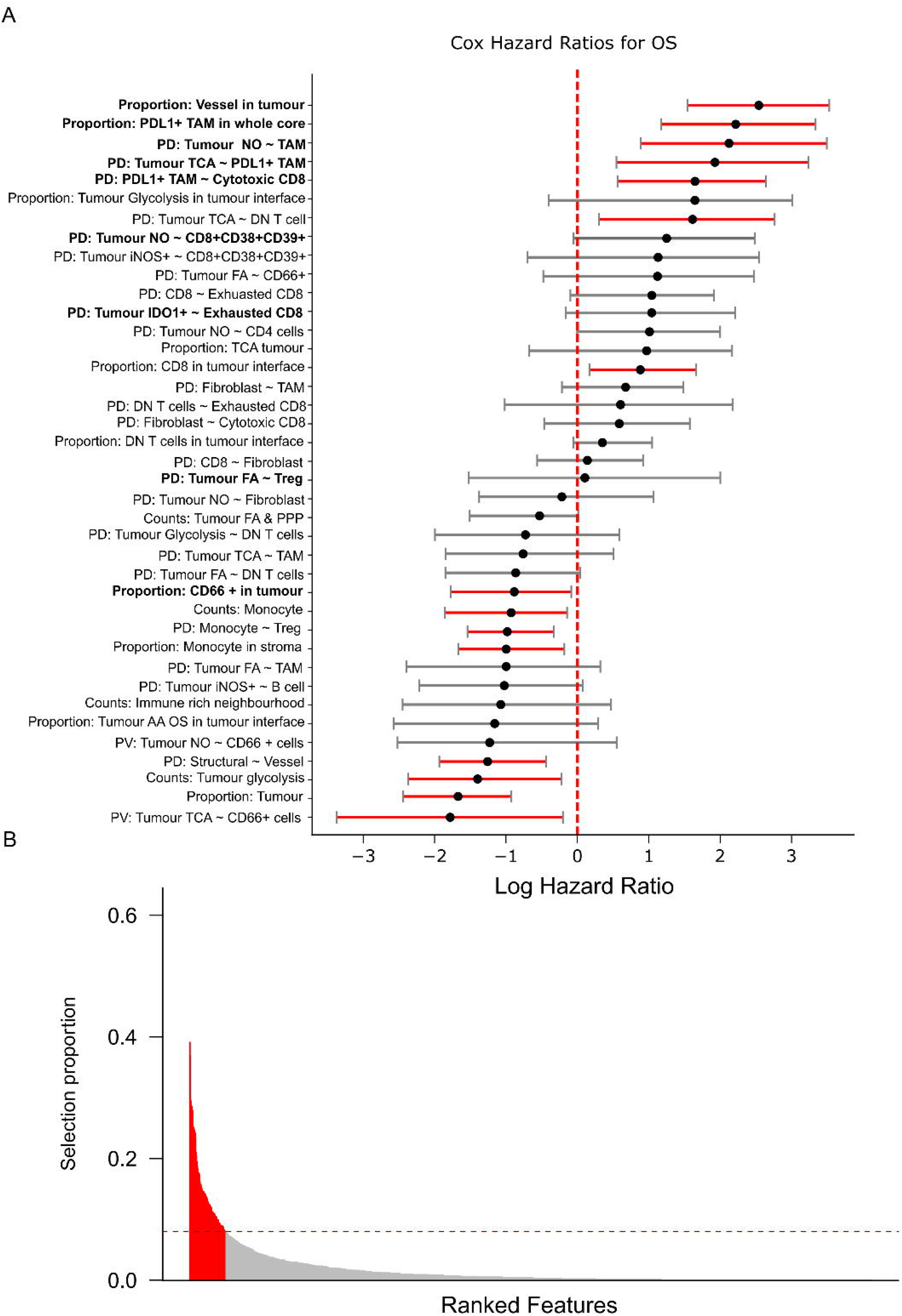
Kaplan Meier survival models for proximity density scores between PDL1+ tumour associated macrophages (TAM) and (**A**) Ki67+, (**B**) iNOS+, (**C**) CD44+, and (**D**) IFN-g +.

**Figure 6:**
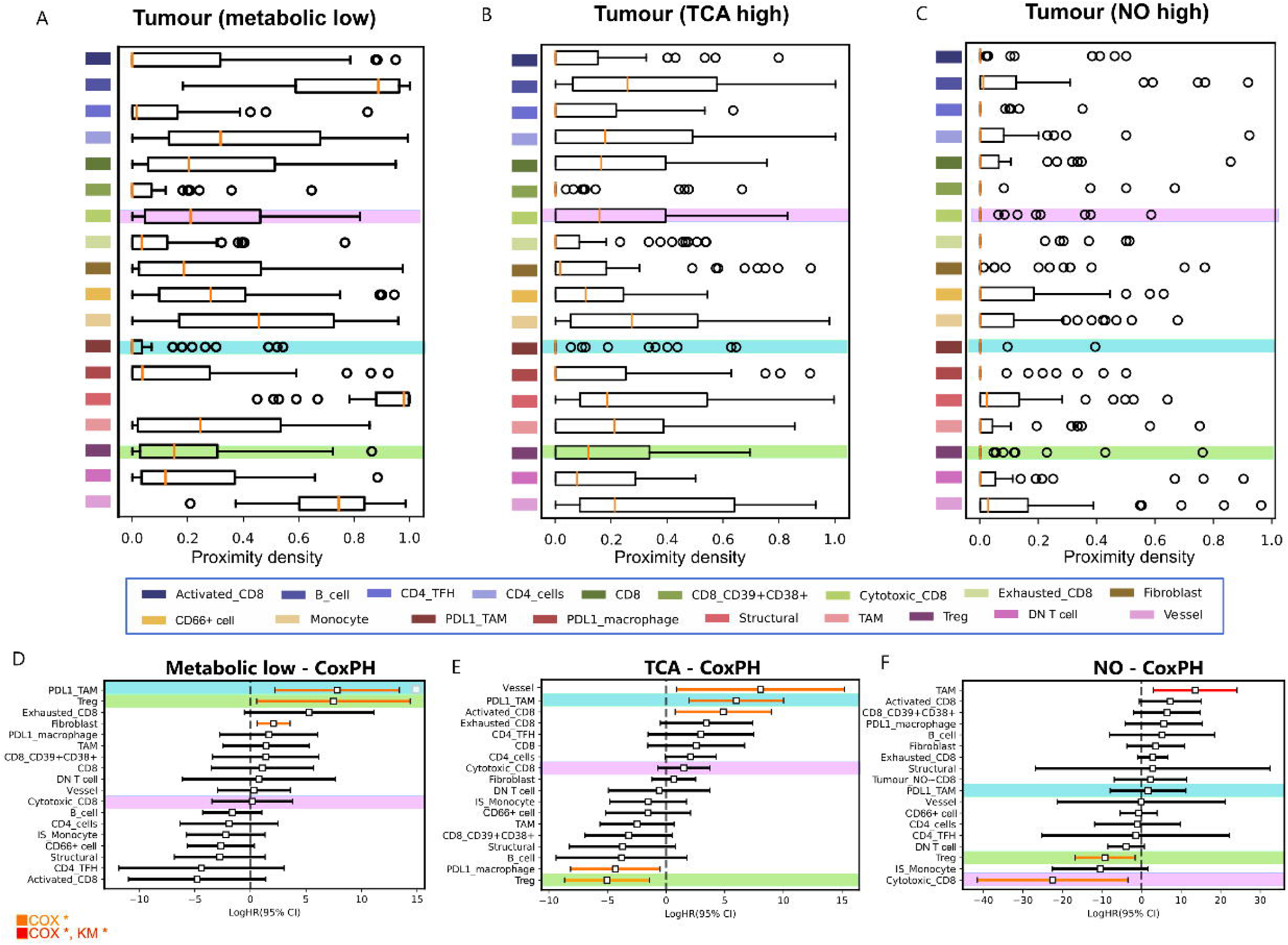
Stability selection results from CoxPH models fit to OS survival data. **A)** Hazard ratios for features with non-zero CoxPH coefficients after selection by a stability selection method. Features that are statistically significant in the univariate are shown in bold and confidence intervals that do not cross Log Hazard Ratio = 0 are shown in red. **B)** Feature selection proportions at the optimally stable selection frequency threshold.

To confirm the validity of our interpretation of the univariate selected features, we implemented a stability-selection method on the full set of considered features, including proportions, cell counts, neighbourhood proportions, proximity density, and proximity volume. The stability selection method bootstrapped and fit CoxPH models to the cohort’s endpoint data, selecting 52 features of interest after a final regularized CoxPH fit. A number of these features, particularly vessel and fibroblast density features, are correlated with disease outcome and were therefore not considered in the univariate analysis. Some features are spatial proximity volume metrics, which were not considered because this metric was less biologically interpretable than proximity density. 9 out of 50 of these features align with the univariate-selected features as interpreted in this analysis. Both the log HR and confidence interval for 15 of these 50 features was either > 0 or < 0. Of these 15, 7 features align with the univariate selected features providing a robust indication that these considered features were selected in a false discovery rate-controlled manner. The HR direction also aligns with the predicted survival endpoints in the univariate analysis for all features other than 1.

## DISCUSSION

HGSOC is the most common type of ovarian cancer, and because of mild or misleading symptoms, it is often diagnosed at a late stage when therapy resistance is common^2^. Understanding the immune contexture of HGSOC in a treatment resistant landscape is important for improving future therapies and clinically overcoming mechanisms of resistance. Ovarian cancer is an immunogenic tumour that exhibits spontaneous antitumour immune response^30^, where the infiltration of lymphocytes into the TME is more often associated with favourable prognosis^30^. However, while some subtypes of these immune cell populations exert anti-tumour activity, other subtypes can promote tumour growth. Here, we used a high-plex spatial proteomic analysis to gain greater insight into the distribution and heterogeneity of immune cells and their functional states thereby to understand how these cells may impact disease progression. This study highlights the existence of location-based patterns among TME cells of HGSOC patients, with a focus on their functional and metabolic states that are associated with overall survival.

Previous studies have shown that tumour infiltrating lymphocytes (TILS) have a prognostic benefit in ovarian cancer, and in particular, the presence of CD8+ T cells in the tumour has been associated with improved survival^31^. CD8+ T cells are considered integral in anti-tumour activity, where they secrete cytotoxic molecules, including Granzyme B to trigger tumour cell apoptosis^31^. Within our cohort, we found that patients who had an increased proportion of cytotoxic CD8+ T cells (characterised by granzyme B positivity) within a 10-µm distance of the dense tumour neighbourhoods had improved survival. When we increased the radius from the tumour, the cytotoxic CD8+ T cells no longer appeared to be prognostic, reasonably indicating that, for effective CD8+ T cell function, these cells may need to be in close proximity to the tumour to have benefit. Tumour infiltrating scores have proven to be a reliable prognostic tool in a range of cancers, however, it has been stated that the spatial organisation of these cells provides additional valuable information^32^. Here, we highlight the importance of location-based metrics for predicting clinical outcomes. Reasons that these cells may not be reaching the tumour, or exerting anti-tumour activity at these close distances may be due to immune evasion mechanisms that have evolved in the tumour, including dysregulated metabolism and expression of immunomodulatory markers^33^. Studies have indicated that the immune evasion mechanisms in ovarian cancer are highly heterogeneous and can by influenced by the metastatic spread and location of tumours^34,35^.

Dysregulated tumour metabolism plays a role in tumour growth and immune evasion^8^. We assessed how the changes in metabolic profile of tumour cells associated with the location of immune cell within the TME and subsequently, how this impacted patient outcomes. In relation to the tumour cells that were categorized as metabolically low, due to not highly expressing any of the metabolic markers in the IF panel, we found that patients who had a higher proportion of their T-Reg cells within a 50µm radius of these tumour cells had a poorer overall survival. This is not surprising as the presence of T-Reg cells in the tumour microenvironment often suppress the activity of the CD8+ T cells by secreting immunosuppressive cytokines^36^. Interestingly, in tumours with high citrate synthase or inducible nitric oxide synthase (iNOS) expression, the presence of Tregs was associated with improved survival, leading us to speculate that the byproducts of these dysregulated metabolic pathways may negatively affect the function of Tregs. Furthermore, when looking at the iNOS tumours, cytotoxic CD8+ T cells were also associated with improved outcomes, which could the result of a favourable TME or Treg dysfunction allowing the cytotoxic CD8 T cells to be more effective. Nitric oxide in high concentrations can be anti-tumorigenic, contributing to cytotoxicity and may have pro-apoptotic effects. Overexpression of iNOS in other cancers, including prostate and colon cancer, have shown an increased sensitivity to cisplatin or radiotherapy. Studies have reported that NO modulates the T cell response and T cell differentiation^37^. For example, it has been shown that exogenous production of NO production is important for cytotoxic CD8 T cells to target and kill tumour cells and can inhibit Treg induction^37^.

The presence of tumour associated macrophages in HGSOC has been found to be negatively associated with overall survival^38^. Depending on the origin of TAMs, they can display M1 or M2 macrophage characteristics. Typically, in ovarian cancer, TAMs are more often of an M2 phenotype and are known to have immunosuppressive properties in the TME by releasing pro-tumour cytokines and chemokines ^38^. Furthermore, the expression of PDL1 by TAMs can directly suppress the activity of cytotoxic CD8 T cells^38^. In our cohort, we identified PDL1+ TAMS to be associated with worse overall survival. Within our dataset, however, we have seen a higher proportion of PDL1 TAMs in a 50 µm radius of tumour cells, activated CD8 T cells, CD8 T cells, cytotoxic CD8 T cells, or exhausted CD8 T cells that are associated with worse overall survival (Supplementary figure 4).

CEACAM1 (CD66a) is a cell adhesion molecule that has been characterised to have both pro-tumour and anti-tumour effects depending on the tumour type. In ovarian cancer, the RNA and protein expression of CEACAM1 has been found to be an independent prognostic marker for improved overall survival^39^. It was also included as one of six markers for a 6-plex assay that aimed to improve and model diagnostics of ovarian cancer^40^. This study found that in plasma, CEACAM1 was higher in patients with benign ovarian tumours compared to patients with cancerous lesions and suggested that CEACAM1 plays a role in tumour suppression ^40^. Aside from the above-mentioned findings, there is currently limited published data available describing CEACAM1 in ovarian cancer. Our data supports CD66 as a positive prognostic marker at a whole core level, and when these cells localise closer to tumour cells, patients have improved survival outcomes. Given that the CD66 antibody used in this panel target CD66a, c and e epitopes, we are unable to definitively support these findings. However, our data does suggest that potentially we are seeing a similar relationship. CD66a, b, c and d epitopes are known to be expressed on neutrophils, but can also be expressed on epithelial, plasma and malignant cells^41,42^. The role of neutrophils in ovarian cancer is controversial, with a study finding that neutrophilia and the presence of granulocyte colony stimulating factor in the TME are linked with chemotherapy and radiotherapy resistance, metastases, immune suppression, and cancer cell stemness^43^. However, neutrophils can undergo polarization to either an anti-tumour phenotype (N1) or a pro-tumour phenotype (N2) depending on the inflammatory state of the TME^44^. The characterisation of N1 and N2 phenotypes using surface markers remains a challenge of the field, and with our antibody panel we were unable to characterise these CD66+ cells further. N1 neutrophils can contribute to anti-tumour immune activity through the excretion of cytotoxic mediators including TNF-α, NO, and H O ^45^. Given that we see iNOS upregulated for our CD66^+^ cells, this indicates a potential anti-tumour effect from this population of cells.

This study provides valuable insight into the TME of HGSOC, however, there are limitations of this study that we acknowledge. One limitation of this cohort is that there was no information available regarding the time until reoccurrence and whether, and potentially which, post-surgery treatments were administered. Differences in treatment regimens will impact patient outcomes and is something that we were unable to control. The differences in preservation of the samples and the dates of collection mean that sample quality varies throughout the TMA. While future studies would benefit from a larger sample size and increasing tumour area captured, this study provides comprehensive mapping and analysis of the tumour microenvironment of HGSOC, allowing insight into the metabolic and functional state of many cells. Furthermore, because the tumour microenvironment is dynamic, temporal analysis of samples would provide better information about the evolution of the tumour microenvironment.

## CONCLUSION

Spatial profiling of HGSOC revealed that patient overall survival may be impacted by dysregulated metabolism of tumour cells and the effects this has on nearby immune cells. Additionally, the function of immune cells could be associated with good or poor survival depending on the main metabolic pathway upregulated in the tumour. Furthermore, the presence of CD66+ cells and PDL1+ tumour associated macrophages appeared to play a significant role in disease outcomes. The results from this study highlight the importance of maintaining the spatial location of cells. This was, particularly evident, where we identified that the distance of cytotoxic CD8 T cells from the tumour boundary, which was significant for overall survival. Our results highlight the importance of spatial profiling as the localisation of immune cells within the TME can indicate whether these immune cells are effective.

## Data Availability

All data produced in the present study are available upon reasonable request to the authors

## Abbreviations

AR9: Antigen retrieval 9
AUC: Area under the curve
GDC: Genomic data commons
FFPE: Formalin fixed paraffin embedded
GMM: Gaussian Mixture Model
GPU: Graphical processing unit
H2O: Water
H2O2: Hydrogen peroxide
HGSOC: High Grade Serous Ovarian Cancer
HLA: Human Leukocyte Antigen
HR: Hazard Ratio
IF: Immunofluorescence
KM: Kaplan Meier
KNN: K-Nearest Neighbours
NO: Nitric Oxide
NOS: Nitric Oxide Synthase
OC: Ovarian Cancer
OS: Overall survival
PH: Proportional Hazard
QC: Quality control
SD: Standard deviation
TAM: Tumour associated macrophage
TCA: Tricarboxylic acid cycle
TCGA: The Cancer Genome Atlas
TILS: Tumour infiltrating lymphocytes
TMA: Tumour microarray
TME: Tumour microenvironment.

## AUTHOR CONTRIBUTIONS

Concept/Design: AK1, JF. Experimentation: NB, MD. Analysis: NB, AK. Writing and critical review: all authors. AK: Aaron Kilgallon; AK1: Arutha Kulasinghe

## ACKNOWLEDGEMENTS

The authors would like to acknowledge the support from the Wesley Research Institute, Frazer Institute (University of Queensland), and the Translational Research Institute.

