## Supplementary for "Proximity and metabolic activity of the Tumour Microenvironment as predictors of survival in High Grade Serous Ovarian Cancer (HGSOC)"

**Supplementary figures**

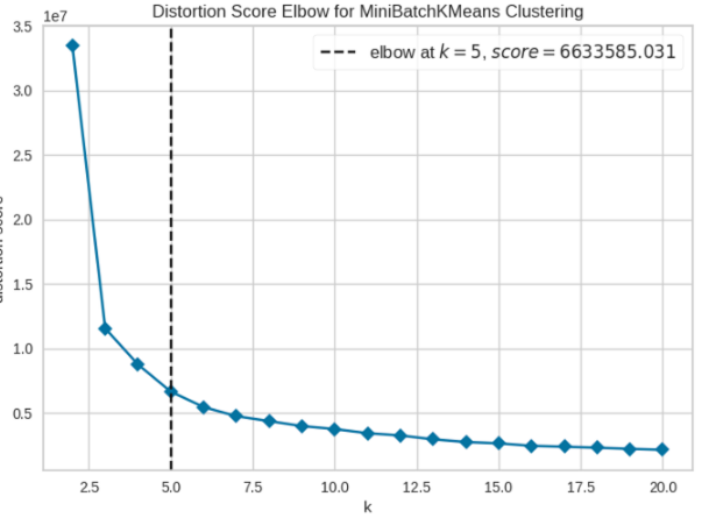

**Supplementary Figure 1:** Distortion score for MiniBatchKMeans Clustering for neighbourhood generation.

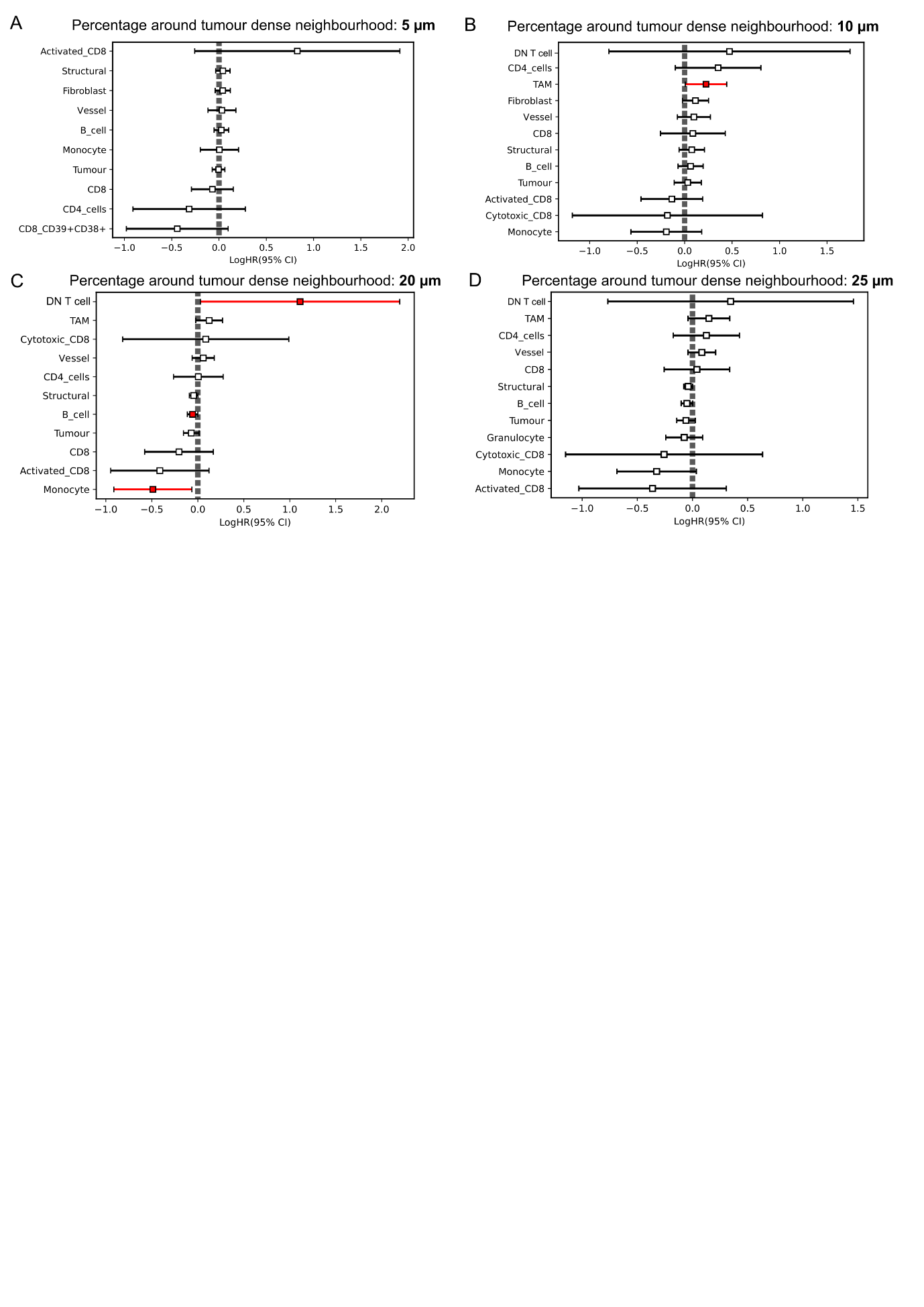

**Supplementary Figure 2:** Cox Proportional Hazard Ratio based on percentage of cell types surrounding tumour dense neighbourhood patches at 5 µm (A), 15 µm (B), 20 µm (C) and 25 µm (D) radii.

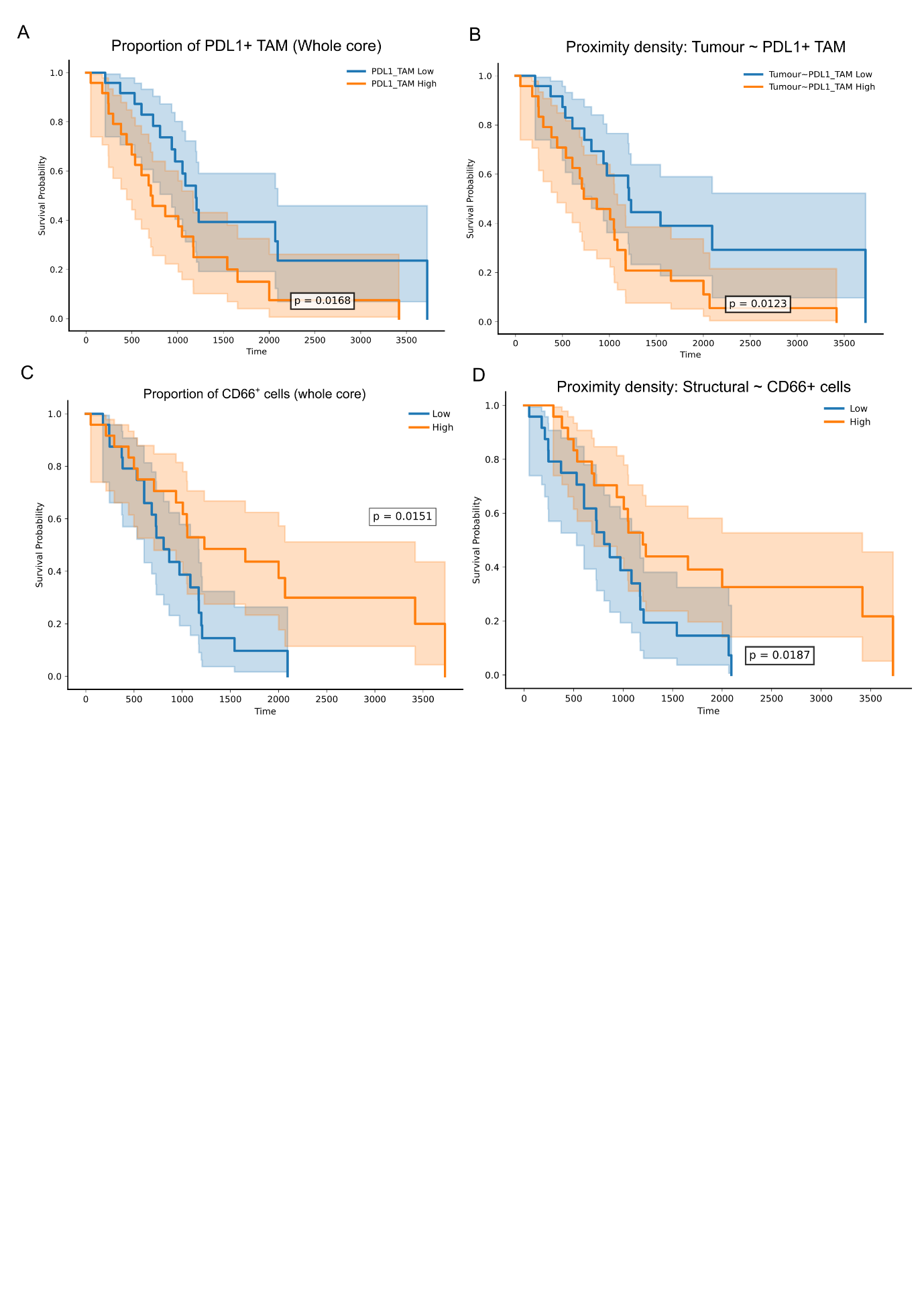

**Supplementary figure 3**: Kaplan Meier (KM) survival analysis, patients stratified based on median score. High scores shown in orange, low scores shown in blue. A) KM survival curves calculated based on the proportion of PDL1+ TAMs in each core. B) KM survival curves calculated based on the proximity density score between tumour cells and PDL1+ TAMs in each core. C) KM survival curves calculated based on the proportion CD66+ cells in each core. B) KM survival curves calculated based on the proximity density score between structural cells and CD66+ cells in each core.

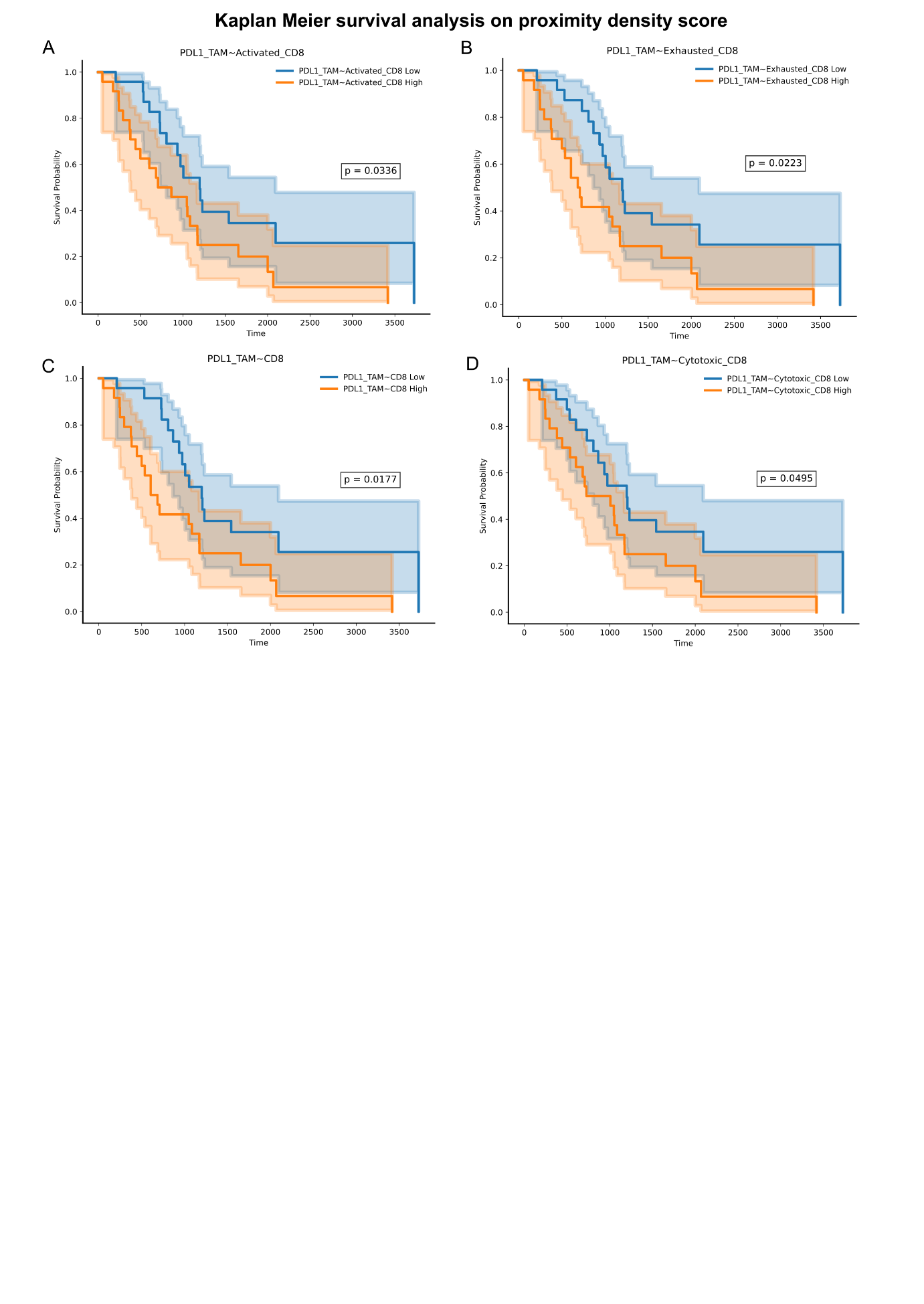

**Supplementary figure 4:** Kaplan Meier survival analysis based on proximity density scores between PDL1+ TAMS and activated CD8 T cells (A), exhausted CD8 T cells (B), CD8 T cells (C), and cytotoxic CD8 T cells (D). Patients were stratified into low (blue) and high (orange) based on the median score of the feature.

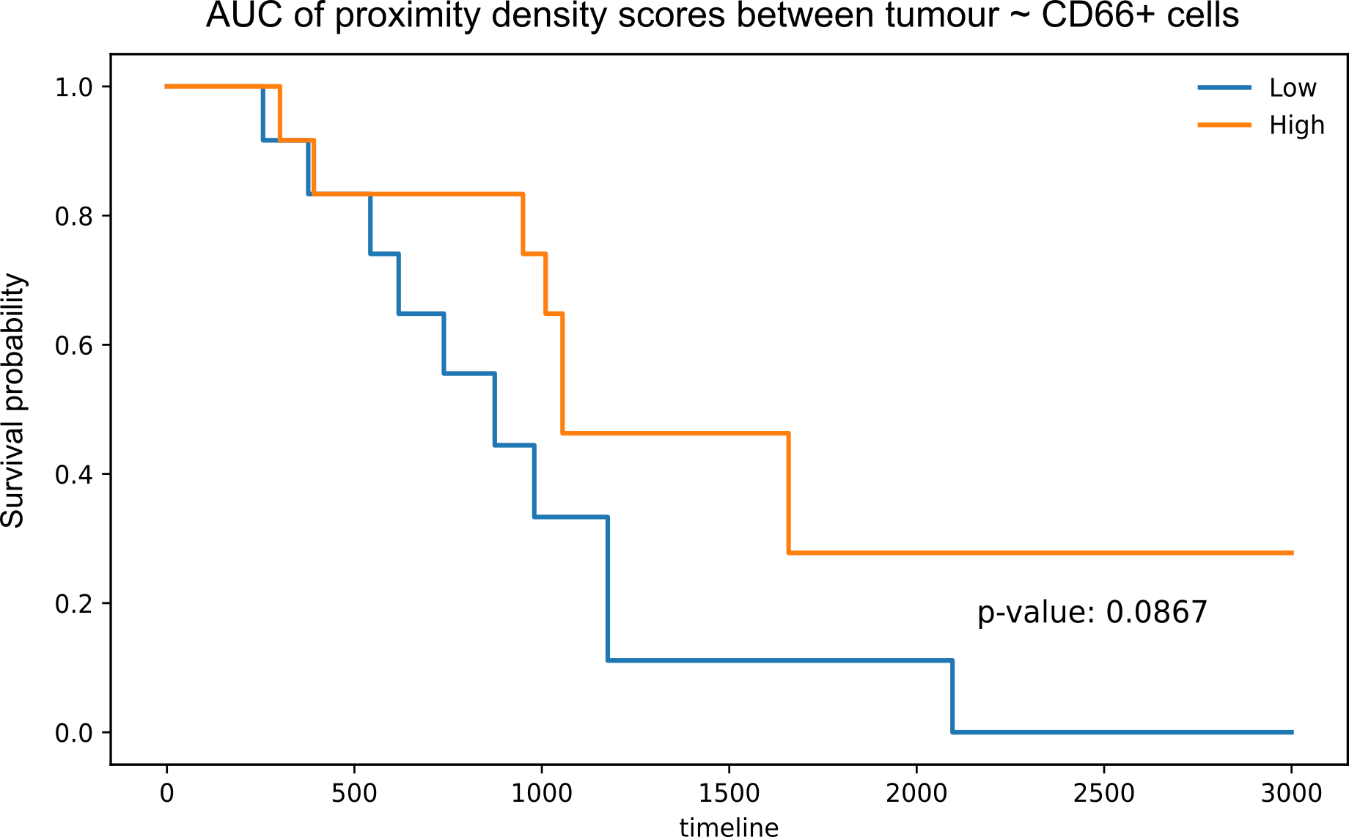
**Supplementary figure 5**: Kaplan Meier analysis based on area under the curve (AUC) scores generated from proximity density scores between tumour cells and CD66+ cells. Proximity density scores were calculated over a range of distances from 25-100 µm at 5 µm increments. Based on AUC for each sample, the upper and lower quartiles were modelled using Kaplan Meier survival analysis and long-rank test for statistical significance.

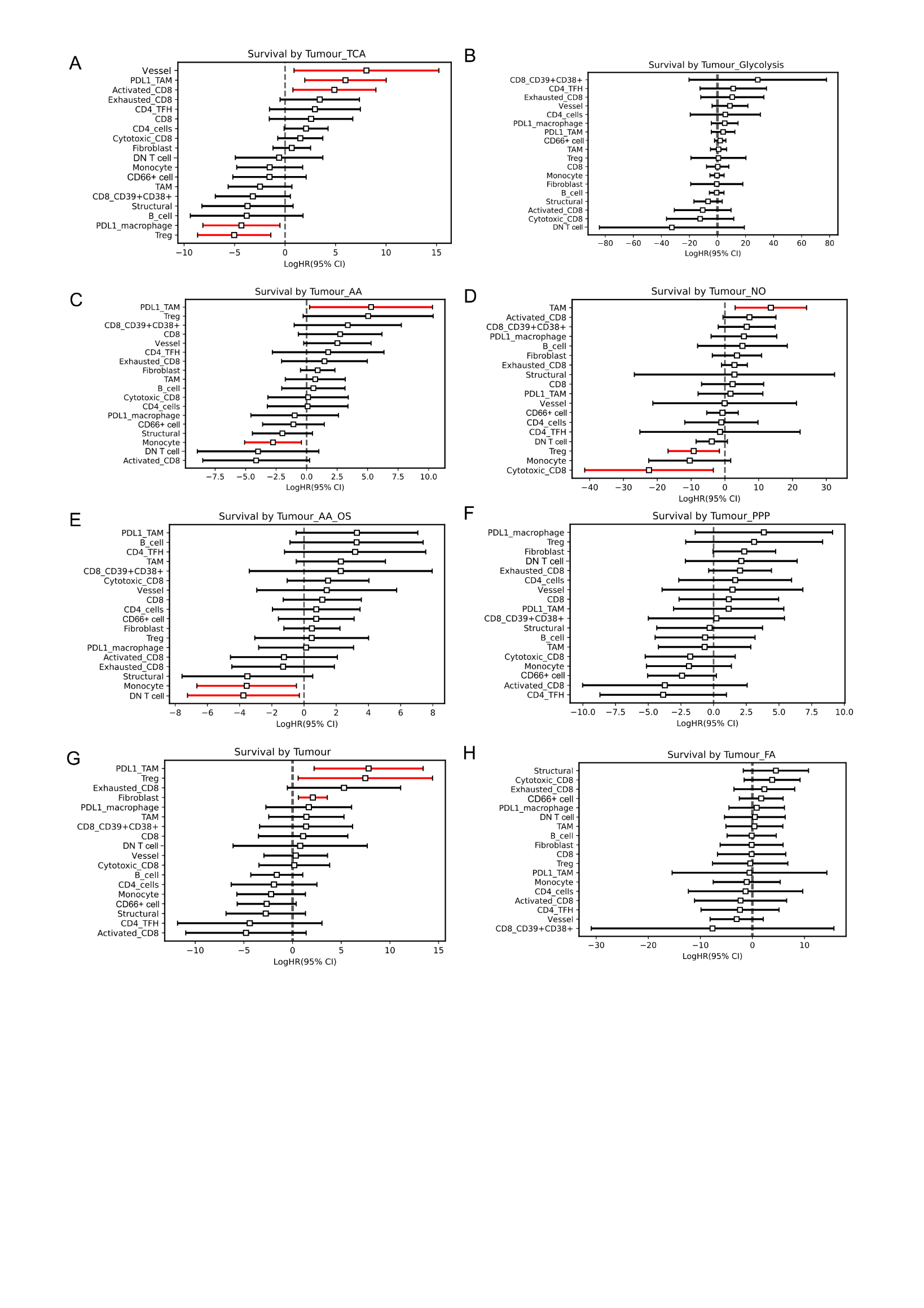
**Supplementary figure 6:** Cox proportional hazard models based on proximity density scores of metabolically active tumour cells with immune cells. Proximity density scores were calculated at a 50 µm radius. Statistically significant findings are highlighted in red.

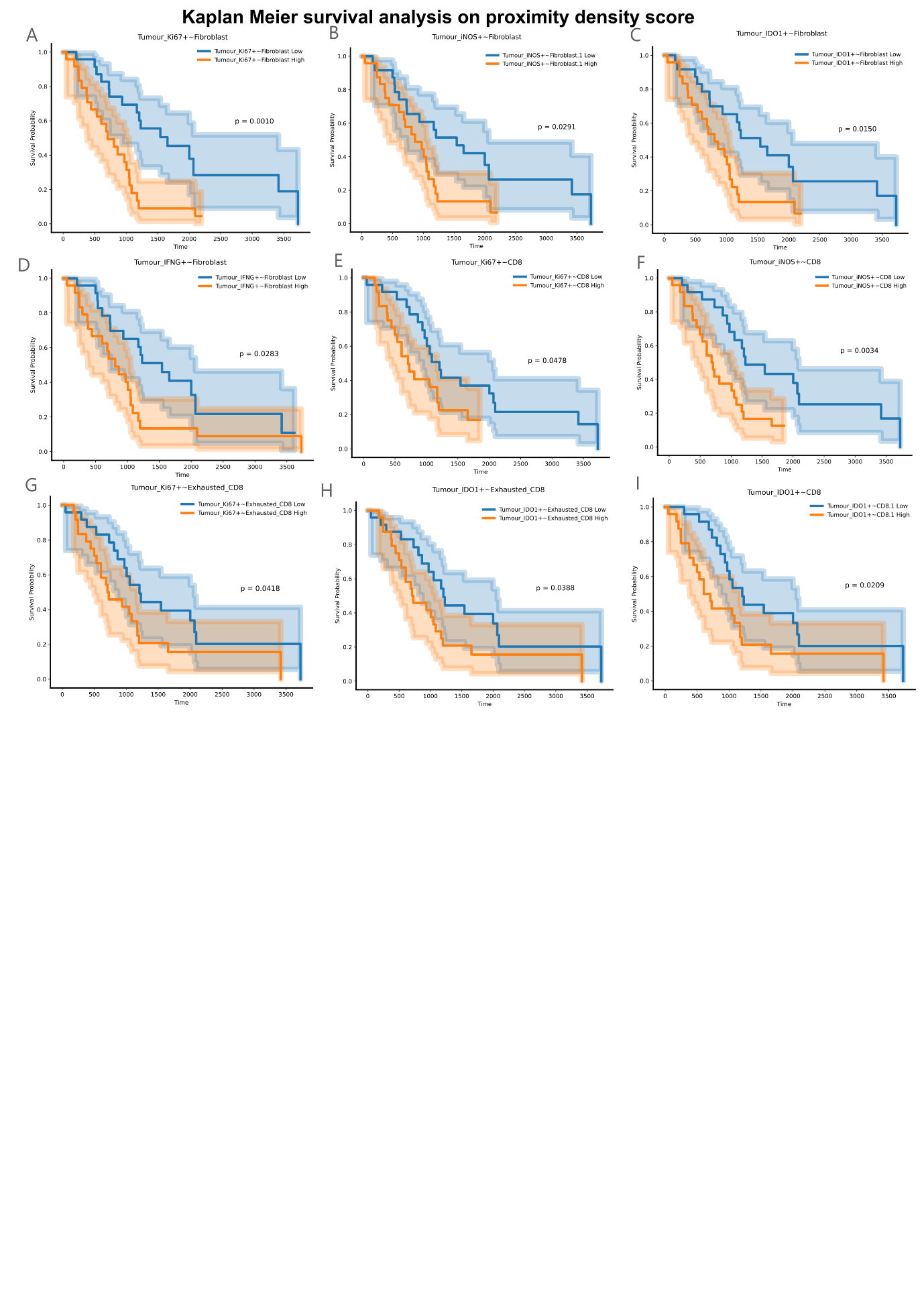
**Supplementary figure 7:** Kaplan Meier survival curves using proximity density scores between functional tumour phenotypes and immune cells. Patients are into high (orange), and low (blue) groups based on the median feature score. Statistical significance is calculated using the log-rank test.
